# Chemosensory dysfunctions induced by COVID-19 can persist up to 7 months: A study of over 700 healthcare workers

**DOI:** 10.1101/2021.06.28.21259639

**Authors:** Nicholas Bussière, Jie Mei, Cindy Lévesque-Boissonneault, Mathieu Blais, Sara Carazo, Francois Gros-Louis, Gaston De Serres, Nicolas Dupré, Johannes Frasnelli

## Abstract

Several studies have revealed either self-reported chemosensory alterations in large groups or objective quantified chemosensory impairments in smaller populations of patients diagnosed with COVID-19. However, due to the great variability in published results regarding COVID-19-induced chemosensory impairments and their follow-up, prognosis for chemosensory functions in patients with such complaints remains unclear. Our objective is to describe the various chemosensory alterations associated with COVID-19 and their prevalence and evolution after infection. A cross-sectional study of 704 healthcare workers with a RT-PCR confirmed SARS-CoV-2 infection between 28/2/2020 and 14/6/2020 was conducted 3 to 7 months after onset of symptoms. Data were collected with an online questionnaire. Outcomes included differences in reported chemosensory self-assessment of olfactory, gustatory, and trigeminal functions across time points and Chemosensory Perception Test scores from an easy-to-use at-home self-administered chemosensory test. Among the 704 participants, 593 (84.2%) were women, the mean (SD) age was 42 (12) years, and the questionnaire was answered on average 4.8 (0.8) months after COVID-19. During COVID-19, a decrease in olfactory, gustatory, and trigeminal sensitivities were reported by 81.3%, 81.5% and 48.0% respectively. Three to seven months later, reduced sensitivity was still reported by 52.0%, 41.9% and 23.3% respectively. Chemosensory Perception Test scores indicate that 19.5% of participants had objective olfactory impairment. These data suggest a significant proportion of COVID-19 cases have persistent chemosensory impairments at 3 to 7 months after their infection but the majority of those who had completely lost their olfactory, gustatory, and trigeminal sensitivity have improved.

## Introduction

Coronavirus disease-2019 (COVID-19) is an ongoing major public health challenge. Olfactory dysfunction (OD) is a specific symptom that may affect approximately 60% of patients suffering from COVID-19 (Spinato, Fabbris et al. 2020, von Bartheld, Hagen et al. 2020, Whitcroft and Hummel 2020), and is now considered as a stronger indicator of COVID-19 than fever, cough and shortness of breath (Gerkin, Ohla et al. 2021).

OD can be quantitative or qualitative. Quantitative OD is defined by a reduction of olfactory sensitivity which can be either a complete (anosmia) or a partial (hyposmia) loss of olfactory function (Hummel, Whitcroft et al. 2016). Qualitative OD describes an altered perception of olfactory stimuli: For example, parosmia is defined as the perception of qualitatively altered smells, and phantosmia is defined as the perception of a smell in the absence of an objective odorant (Hummel, Whitcroft et al. 2016, Sjölund, Larsson et al. 2017). Overall, the prevalence of OD in the general population is around 20% (Landis, Konnerth et al. 2004, Yang and Pinto 2016), and all different forms of OD are associated with reduced quality of life (Croy, Nordin et al. 2014). In addition to OD, COVID-19 also appears to affect other chemosensory modalities, i.e., gustation and trigeminal function (Cooper, Brann et al. 2020, Parma, Ohla et al. 2020).

Olfactory and other chemosensory dysfunctions may have detrimental effects. First, affected individuals can expose themselves to harmful substances such as smoke, gas or spoiled food (Gonzales and Cook 2007, Schiffman 2007). It may trigger dysfunctional nutritional patterns like increased salt and sugar consumption, or anorexia (Mattes, Cowart et al. 1990, Aschenbrenner, Hummel et al. 2008). Individuals with OD also have higher rates of anxiety and depression (Croy, Nordin et al. 2014, Kohli, Soler et al. 2016). Moreover, a functioning olfactory system may be a necessity in some workplaces, such as healthcare, where staff are required to have the ability to detect and qualify the smell of urine, excrement, infected wounds or abnormal smells of breath (Kelly 2012).

Investigation of the long-term effects of COVID-19 on chemosensory function is hindered by the recent onset of the pandemic and other challenges: First, many studies on the prevalence of OD during COVID include a relatively small number of participants (Hintschich, Wenzel et al. 2020, Le Bon, Pisarski et al. 2020) or participants with severe forms of COVID-19 (Moein, Hashemian et al. 2020, Speth, Singer-Cornelius et al. 2020). Secondly, many studies on the prevalence of OD during COVID-19 also include participants with an unclear diagnosis of COVID-19, and/or self-diagnosis (Hopkins, Surda et al. 2020, Parma, Ohla et al. 2020). Lastly, while individuals with anosmia can usually evaluate their olfactory function with accuracy (Lötsch and Hummel 2019), this self-assessment is often challenging for individuals with intermediate forms of OD (e.g., hyposmia) (Landis, Hummel et al. 2003). Finally, studies on persistent post-COVID-19 OD in the past year have used various designs (objective measures (Lechien, Chiesa-Estomba et al. 2021), semi-objective (Petrocelli, Cutrupi et al. 2021), or self-reported (Havervall, Rosell et al. 2021, Hopkins, Surda et al. 2021) and collected data at varying time intervals after onset of disease. For these reasons, to this date, no consensus has been reached regarding the prevalence of post-COVID-19 OD (Xydakis, Albers et al.).

To comprehensively understand long-term olfactory, gustatory, and trigeminal alterations after COVID-19, we analyzed questionnaire responses from a cohort of healthcare workers infected with SARS-CoV-2 during the first wave of the pandemic (February - June 2020). We also developed a Chemosensory Perception Test (CPT), a formal test employing common household odorants and tastants to enable accessible yet accurate self-evaluation of chemosensory functions remotely on a large scale. The CPT is particularly useful when in-person testing is unsafe and testing a large group of participants at distance with mailable tests such as the UPSIT (Doty, Shaman et al. 1984) is costly. Moreover, distance testing has been reported to accurately monitor disease progression in at risk populations (Vaira, Hopkins et al. 2020, Weiss, Attuquayefio et al. 2020).

## Materials and Methods

### Participants

Participants were recruited from a Quebec healthcare worker cohort who have had SARS-CoV-2 infection between 28/2/2020 and 14/6/2020. They were part of a study from the Institut National de Santé Publique du Québec and had agreed to be contacted for other research projects(Carazo 2021). Inclusion criteria were (1) RT-PCR confirmed COVID-19 (2) above 18 years of age, (3) French or English speakers, (4) completed the online questionnaire, and (5) did not report of other respiratory diseases (bacterial or viral infection, or/and allergies with rhinorrhea) within 2 weeks prior to questionnaire completion or chronic sinusitis (Figure 1).

**Figure 1.**
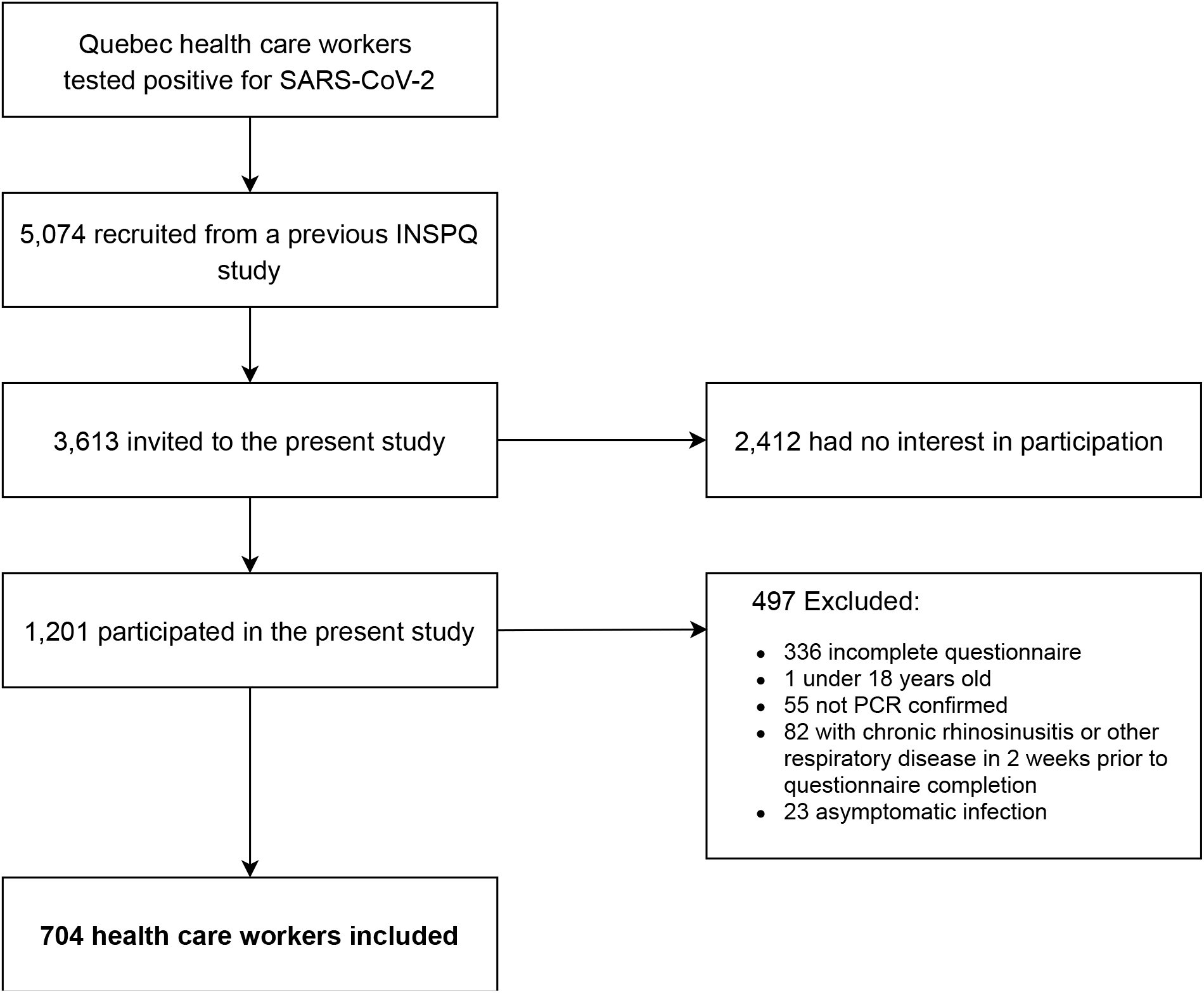
Flowchart of participant inclusion/exclusion procedures. Flowchart of the study design. INSPQ: Institut national de santé publique.

This study was reviewed and approved by the research ethics board of the CHU de Québec – Université Laval (MP-20-2021-5228) and all protocols were reviewed by an independent Scientific Review Committee. This study also complies with the Declaration of Helsinki for Medical Research Involving Human Subjects. All participants provided an online informed consent prior to participation. The study received funding from the Fonds de recherche du Québec-Santé. No compensation or incentive was offered for participation. Data were collected from August 11 to October 29, 2020. Up to four attempts were made to reach by email potential participants. At the time of data collection, participants were 3-7 months after the onset of COVID-19 symptoms.

### Online questionnaire

All participants were asked to complete an online questionnaire which was adapted from the core questionnaire of the Global Consortium on Chemosensory Research(Parma, Ohla et al. 2020).

#### Demographic information

In the first part of the questionnaire, demographic information was collected from all participants. Participants were then instructed to provide medical history and indicate the presence of specific COVID-19 symptoms (Figure 2).

**Figure 2.**
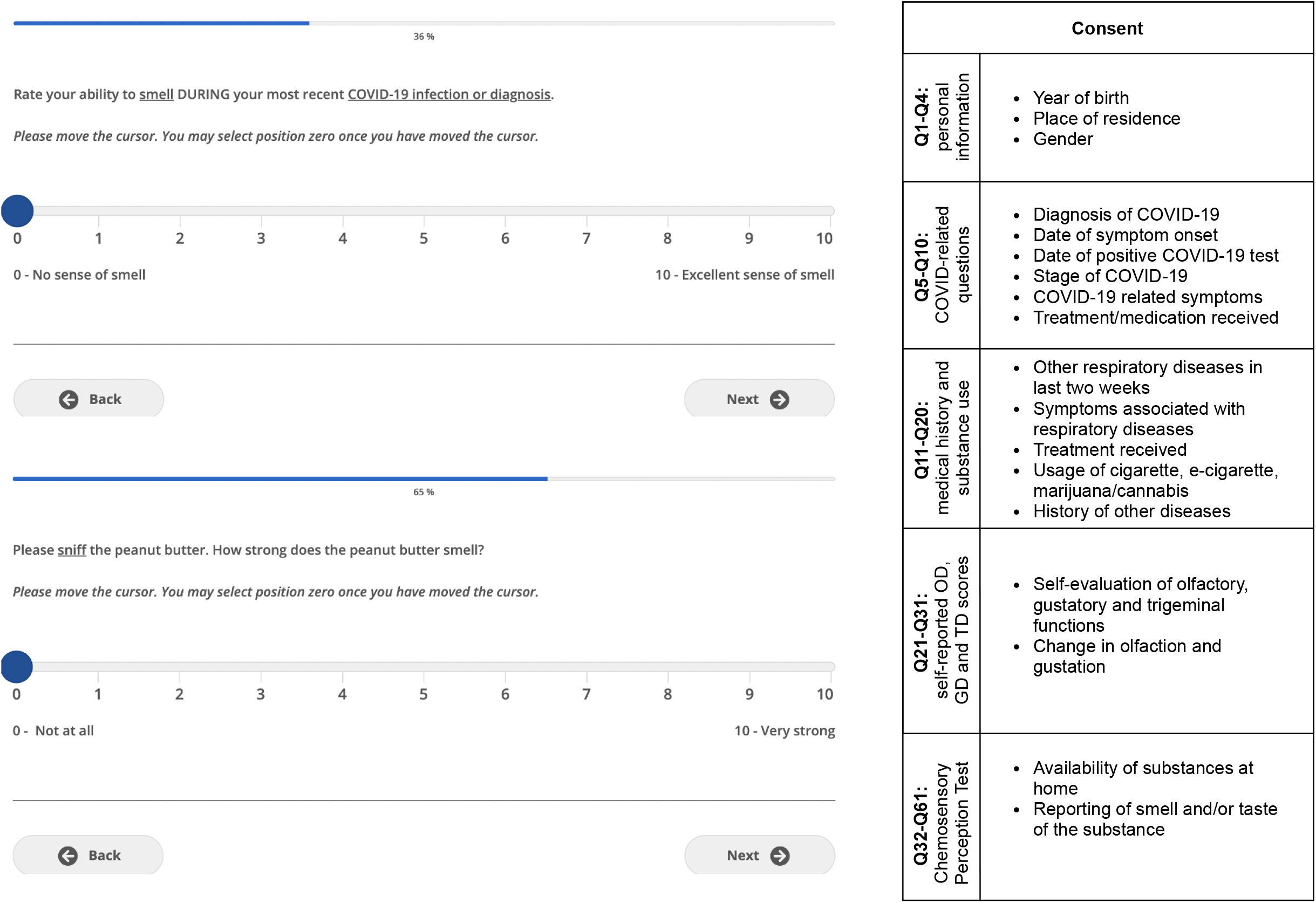
Web-based interface and structure of the online questionnaire. Left, Self-rating of olfaction and reporting of CPT using VAS through the web-based interface, as viewed by the participant. Right, Sections of the online questionnaire. VAS: visual analog scale.

#### Chemosensory self-assessment

Participants were asked to self-evaluate and report their olfactory, gustatory, and trigeminal sensitivity using a 10-point visual analog scale (VAS; Figure 2) for three timepoints: (1) before SARS-CoV-2 infection, (2) during SARS-CoV-2 infection and (3) at questionnaire completion. The specific definition of each chemosensory modality was presented prior to self-evaluation of each chemosensory modality as follows: Olfaction: *The following questions relate to your sense of smell (for example, sniffing flowers or soap, or smelling garbage) but not the flavor of food in your mouth*; Gustation: *The following questions are related to your sense of taste. For example, sweetness, sourness, saltiness, bitterness experienced in the mouth;* Trigeminal: *The following questions are related to other sensations in your mouth, like burning, cooling, or tingling. For example, chili peppers, mint gum or candy, or carbonation*. Further, information on the presence of parosmia or phantosmia following the infection (Landis, Frasnelli et al. 2010) and alterations in the 5 tastes (sweet, salty, sour, bitter, umami) was collected.

#### Chemosensory Perception Test (CPT)

Items commonly found in North American households were used to assess participants’ olfactory and gustatory functions, as odor intensity is the best single predictor to classify individuals with normosmia (Parma, Hannum et al. 2021). Participants had to smell three substances (peanut butter, jam/jelly, and coffee) and rate odor intensity on a 10-point VAS (0: no smell at all; 10: very strong smell). We obtained olfactory scores by averaging these ratings. Pilot data on a total of 93 participants show these scores to accurately detect OD when compared to the Sniffin’ Sticks (cut-off score: 6/10; sensitivity: 0.765; specificity: 0.895; Supplement 3). Participants were asked to prepare saline and sweet water by dissolving respectively a teaspoon of salt or 3 teaspoons of sugar in a cup (250 mL) of lukewarm water. Then, they were asked to taste saline and sweet water and to rate taste intensities on a 10-point VAS. We obtained gustatory scores by averaging these ratings. An ongoing study is comparing CPT gustatory scores with the Waterless-Empirical Taste Test - Self-Administered (Doty, Wylie et al. 2021), but too few participants have been recruited to this to establish its accuracy (Supplement 3).

### Statistical Analyses

A Python script (Python 3.7.5, Python Software Foundation, https://www.python.org) was used to process raw questionnaire data and to calculate the number of participants reporting COVID-19 symptoms, chronic conditions and recent respiratory illnesses. Processed data were analyzed and visualized with SPSS 26.0 (Armonk, NY: IBM Corp), GraphPad Prism 8.3.1 (GraphPad Prism Software, San Diego, CA) and Raincloud plots(Allen, Poggiali et al. 2021).

Parametric (ANOVA) or non-parametric (Friedman) tests were chosen depending on whether normality assumption was fulfilled. To evaluate the effects of COVID-19 on *modality* (olfactory, gustatory, and trigeminal) and *time* (prior to, during and after COVID-19 infection), for *gender* (women, men), repeated measures (rm) ANOVA with age as a covariate were computed. To disentangle interactions, separate rmANOVA were carried out for individual modalities and timepoints with the same factors. Greenhouse-Geisser corrections were used for sphericity and Tukey’s multiple comparisons test were used for post-hoc comparisons. Friedman’s test was followed by Dunn’s post-hoc test to correct for multiple comparisons. To assess the correlation between self-reported olfactory, gustatory, and trigeminal abilities and results of the CPT, Pearson correlation coefficient or Spearman’s rank correlation coefficient was used. For all statistical tests, alpha was set at 0.05. All results are expressed as mean (SD) unless otherwise specified.

## Results

### Characteristics of participants

A total of 704 healthcare workers (593 (84.2%) women, mean age of 42.0 (SD:11.7, range 18 – 70) years were included. The questionnaire was completed on average 4.8 (SD: 0.8, range 3-7) months after symptoms onset. COVID-19 symptoms reported by the 704 participants are listed in Table 1.

**Table 1.** COVID-19 symptoms of the 704 participants.

| Symptoms at time of SARS-CoV-2 infection | No. (%) |
| --- | --- |
| Fever | 353 (50.1%) |
| Dry cough | 361 (51.7%) |
| Cough with mucus | 77 (10.9%) |
| Dyspnea | 316 (44.9%) |
| Chest tightness | 201 (28.6%) |
| Runny nose | 226 (32.1%) |
| Sore throat | 330 (46.9%) |
| Changes in food flavor | 471 (66.9%) |
| Changes in smell | 520 (73.9%) |
| Loss of appetite | 323 (45.9%) |
| Headache | 518 (73.6%) |
| Muscle aches | 444 (63.1%) |
| Fatigue | 611 (86.8%) |
| Diarrhea | 259 (36.8%) |
| Abdominal pain | 102 (14.5%) |
| Nausea | 179 (25.4%) |

### Quantitative disorders

Before COVID-19, average self-reported score was 9.0 (1.6), 9.2 (1.3) and 8.9 (1.9) of 10 for olfaction, gustation and trigeminal function, respectively. Among participants, 0.9%, 0.7% and 1.8% respectively reported an absence of olfaction, gustation and trigeminal function (score 0; Figure 3). During COVID-19, average self-reported score was 2.6 (3.6) for olfaction, 3.4 (3.6) for gustation, and 7.0 (3.0) for trigeminal sensitivity. In the 704 participants, 51.1%, 33.5% and 5.7% reported absence of olfaction, gustation and trigeminal function. At time of questionnaire completion, mean scores were 7.4 (2.5), 8.0 (2.2) and 8.5 (2.2) for olfaction, gustation and trigeminal function respectively and absence of chemical senses was reported respectively by 1.4%, 0.7% and 2.3%. Weak correlations were found between the time since infection and the self-reported olfactory and gustatory scores at questionnaire completion (olfaction: ρ=0.11; gustation: ρ=0.14; both P<.001; trigeminal ρ = .06; P=.11).

**Figure 3.**
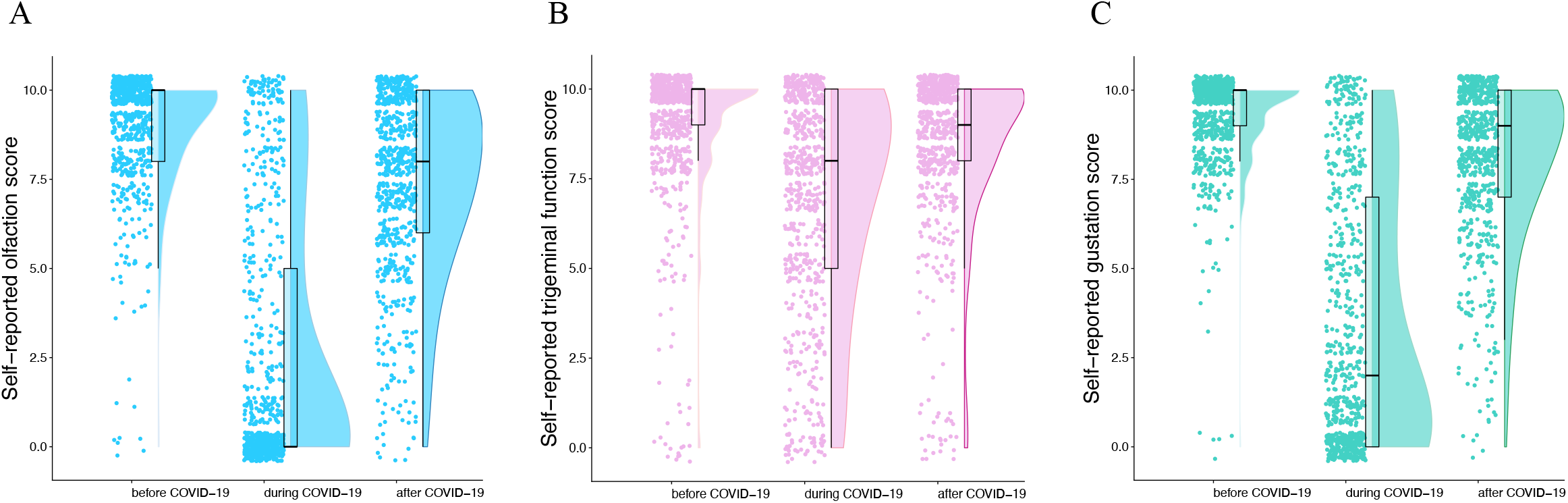
Self-reported scores for the chemosensory modalities before, during and after COVID-19 infection (n=704). Raincloud plot representing self-reported scores for olfaction, gustation, and trigeminal function before, during and after COVID-19. Ratings from individual participants are displayed as dots. Boxplots show the first to third quartiles, horizontal line denotes the median, and whiskers denote 1.5 times interquartile range. Compared to baseline, self-reported scores of olfaction, gustation and trigeminal function were significantly lower during COVID-19 and have not fully returned to baseline values 5 months after COVID-19.

Compared to the baseline chemosensory functions before COVID-19, 572 (81.3%), 574 (81.5) and 338 (48.0%) reported lower olfactory, gustatory, and trigeminal sensitivity during COVID-19. Olfactory and gustatory dysfunction were present in similar proportions (χ^2^(2, N=704) =0.02, *P*=.891) and were different to trigeminal (olfaction: χ^2^(2, N=704) = 174.81 *P*<.001; gustation: χ^2^(2, N=704) = 174.56 *P*<.001). Three to seven months after the infection, 366 (52.0%), 295 (41.9%), 164 (23.3%) reported lower olfactory, gustatory, and trigeminal sensitivity compared to before COVID-19 (Table 2). These proportions were significantly different between all three chemosensory systems (χ^2^(2, N=704) = 123.46, *P*<.001). Overall, there were significant effects of *modality* (F(2,1402)=42.83, *P*<.001, 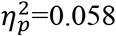; olfactory<gustatory<trigeminal; all *P*<.001), *time* (F(2,1402)=118.47, *P*<.001, 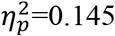; during<after<before; all *P*<.001), and *gender* (F(1,701)=5.52, *P*=0.019, 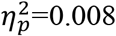; women<men) and significant interactions between these factors (*modality*time, modality*time*gender*; all *P*<.001) on chemosensory self-evaluation. To disentangle these interactions, we analyzed data separately per chemosensory modality and time points.

**Table 2.** Self-reported chemosensory alterations by age group and gender during and 3 to 7 months following COVID-19 (n=704).

|  |  | During acute COVID-19 |  |  | 3-7 months after COVID-19 |  |  |
| --- | --- | --- | --- | --- | --- | --- | --- |
|  |  | Olfaction | Gustation | Trigeminal | Olfaction | Gustation | Trigeminal |
| Age | Gender | N (%) | N (%) | N (%) | N (%) | N (%) | N (%) |
| 18-29 | M (N= 11) | 9 (81.8) | 9 (81.8) | 4 (36.4) | 6 (54.5) | 5 (45.5) | 3 (27.3) |
|  | F (N=115) | 106 (92.2) | 106 (92.2) | 54 (47.0) | 68 (59.1) | 55 (47.8) | 16 (13.9) |
| 30-39 | M (N=26) | 23 (88.5) | 21 (80.8) | 12 (46.2) | 13 (50.0) | 7 (26.9) | 4 (15.4) |
|  | F (N=153) | 133 (86.9) | 129 (84.3) | 77 (50.3) | 83 (54.2) | 63 (41.2) | 37 (24.2) |
| 40-49 | M (N=33) | 23 (69.7) | 23 (69.7) | 13 (39.4) | 12 (36.4) | 9 (27.3) | 5 (15.2) |
|  | F (N=165) | 142 (85.5) | 137 (83.0) | 83 (50.3) | 97 (58.8) | 77 (46.7) | 42 (25.5) |
| 50-59 | M (N=28) | 13 (46.4) | 14 (50.0) | 9 (32.1) | 7 (25.0) | 7 (25.0) | 4 (14.3) |
|  | F (N=128) | 100 (78.1) | 102 (79.7) | 66 (51.6) | 62 (48.4) | 57 (44.5) | 39 (30.5) |
| 60+ | M (N=13) | 5 (38.5) | 6 (46.2) | 5 (38.5) | 2 (15.4) | 1 (7.7) | 3 (23.1) |
|  | F (N=32) | 19 (59.4) | 27 (84.4) | 15 (46.9) | 16 (50.0) | 14 (43.8) | 11 (34.4) |
| **Total (N=704)** |  | **572 (81.3)** | **574 (81.5)** | **338 (48.0)** | **366 (52.0)** | **295 (41.9)** | **164 (23.2)** |

#### Chemosensory modality

With regards to <u>olfactory</u> function, significant main effects of *time* (F(2,1402)=165.07, *P*<.001 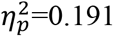; during<after<before; all *P*<.001; Figure 3A), *age* (F(1,701)=4.42, *P*=.012, 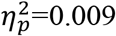) and *gender* (F(1,701)=4.42, P=.036, 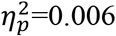; women < men) were revealed. In addition, we observed significant interactions of *time*age* (F(2,1402)=23.39, *P*<.001, 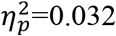) and *time*gender* (F(2, 1402)=21.69, *P*<0.001, 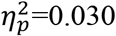). With regards to <u>gustatory</u> function, we observed significant main effects of *time* (F(2,1402)=102.97, *P*<.001, 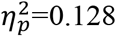; during<after<before; all *P*<.001; Figure 3B) and *gender* (F(1, 701)=9.80, *P*=.002, 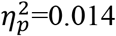; women<men), but no effect of *age*. We also observed significant interactions of *time*age* (F(2, 1402))=5.97, *P*=.005, 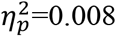) and *time*gender* (F(2, 1402))=20.02, *P*<.001, 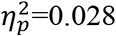).

With regards to <u>trigeminal</u> function, we observed significant main effects of *time* (F(2,1402)=3.91, *P*=.020, 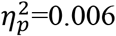; during<after<before; all *P*<.001; Figure 3C), and age ((1,701)=5.08, *P*=.025, 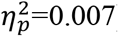) but no effect of *gender*. We also identified significant interactions of *time*age* (F(2, 1402)=4.70, *P*=.016, 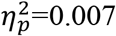) and *time*gender* (F(2, 1402)=4.50, *P*=.019, 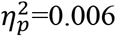).

#### Time point

With regards to chemosensory function <u>before</u> infection, we observed a significant effect of *gender* (F(1,701)=8.52, *P*=.004, 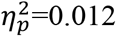; men < women), but not of *modality, age* nor interactions. <u>During</u> COVID-19, we observed a significant effects of *modality* (F(2, 1402)=96.714, *P*<.001, 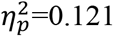; olfaction<gustation<trigeminal; all *P*<.001), *gender* (F(1, 701)=21.98, *P*<.001, 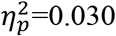; women<men), and age (F(1, 701)=4.74, *P*=.030, 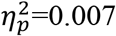). Further, we found significant interactions *modality*\**age* (F(2, 1402)=24.185, *P*<.001, 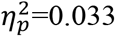) and *modality*gender* (F(2, 1402)=6.76, *P*=.002, 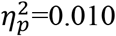). Finally, <u>after</u> infection, we observed a significant effect of *modality* (F(2, 1402)=9.91, *P*<.001, 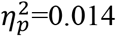; olfaction<gustation<trigeminal; all *P*<.015), but not of *gender* or *age*, nor any interaction.

Compared to baseline (before infection), changes in chemosensory function were correlated for all modalities during infection (olfaction-gustation: ρ=0.69; gustation-trigeminal: ρ=0.43; olfaction-trigeminal: ρ=0.33; all *P*<.001, Figure 4 A-C) and after infection (olfaction-gustation: ρ=0.69; gustation-trigeminal: ρ=0.40; olfaction-trigeminal: ρ=0.36; all *P*<.001, Figure 4 D-F).

**Figure 4.**
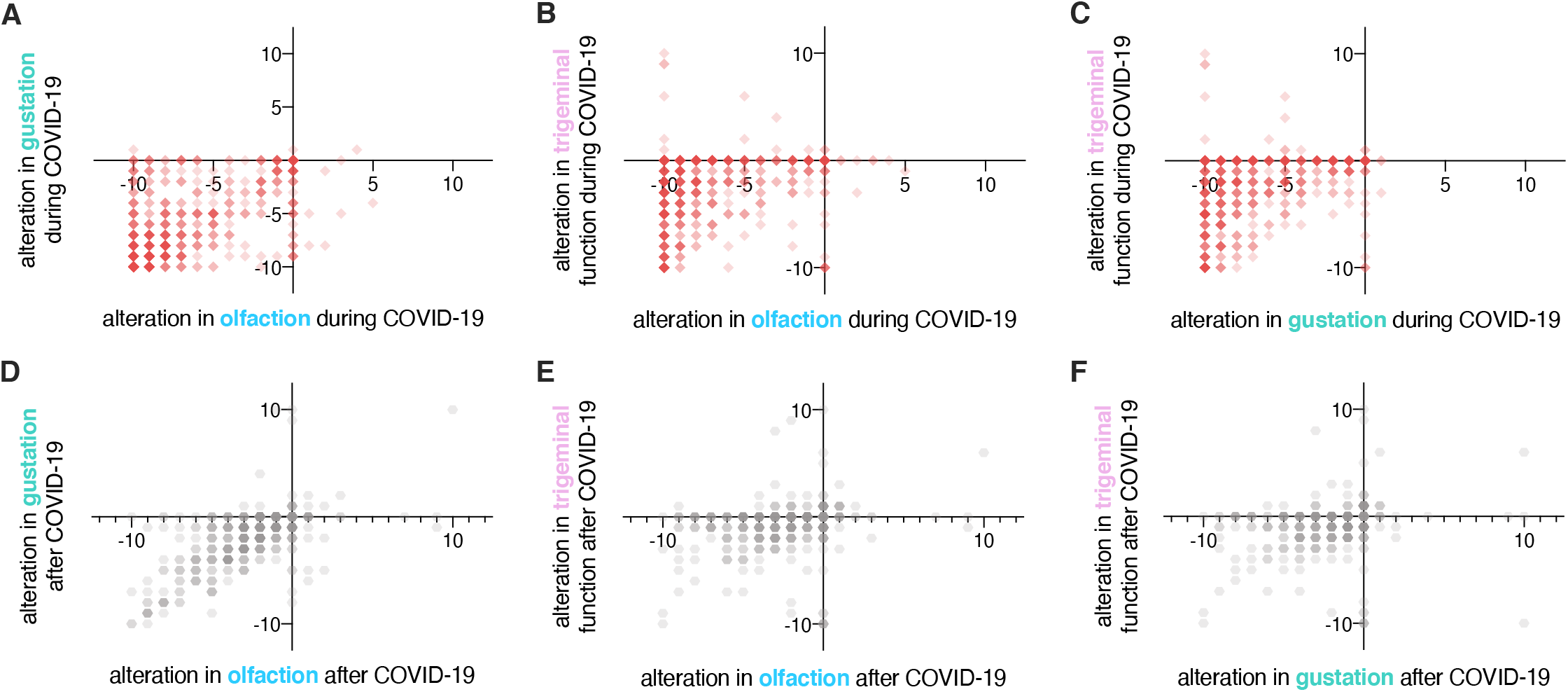
Correlations between alterations in chemosensory modalities (n=704). Red squares, correlations between alterations in olfaction, gustation, and trigeminal functions during COVID-19. Grey hexagons, correlations between alterations in olfaction, gustation, and trigeminal functions after COVID-19. Darker colors indicate higher occurrence.

### Qualitative disorders

Among included participants, 78 (11.1%) reported parosmia, 73 (10.4%) experienced phantosmia and/or 82 (11.6%) had waxing and waning of olfaction following infection. In addition, 42 (6.0%) claimed that they experienced other forms of OD (hyposmia to specific substances, hyperosmia, parosmia only at high concentrations or slow identification times). Furthermore, 335 (47.6%) participants reported changes to perception of sweet, 338 (48.0%) salty, 293 (41.6%) sour, 309 (43.9%) bitter and 281 (39.9%) umami. A total of 275 (39.1%) participants reported alterations in all 5 tastes.

### Chemosensory Perception Test

Among the 704 participants, 137 (19.5%) had a CPT score suggestive of OD. Mean CPT scores were lower for olfaction than gustation (7.84 (1.78) vs 8.42 (2.31); Z=8.193, *P*<.001). Neither *age* nor *gender* had an effect on CPT scores. CPT scores correlated with self-reported chemosensory abilities at testing time (olfaction: ρ=0.67; gustation: ρ=0.51; *P*<.001 for both).

## Discussion

This study reports chemosensory dysfunction 3 to 7 months following SARS-CoV-2 infection in a large cohort of RT-PCR-confirmed healthcare workers. In addition to confirming the now well-established detrimental effect of acute COVID-19 on all three chemosensory systems (olfactory, gustatory, trigeminal), our major findings are: (1) the detrimental effect of COVID-19 lasts beyond the acute phase after the infection, half of those affected indicated that olfactory function had not returned to the baseline levels 3 to 7 months later, while 20% of infected participants reported scores in a formal test that are consistent with the presentation of hyposmia/anosmia; (2) approximately 10% of the patients exhibit parosmia and/or phantosmia; (3) women are more heavily affected than men.

We observed chemosensory dysfunction in the acute phase of COVID-19, which was most pronounced for olfactory function, but less so for gustatory function and even less for trigeminal function. The proportion of participants describing OD and GD in the acute phase of COVID-19 in this study was comparable to earlier studies (Hajikhani, Calcagno et al. 2020). Although the proportions of participants indicating a decrease in olfaction or gustation were comparable, the olfactory system seems to be more severely impaired. Given the cross-sectional design of the present study, recall bias may have a role to play in the prevalence of OD and GD in similar study populations, but published studies with little to no recall bias also report equivalent prevalence of OD and GD (Andrews, Pendolino et al. 2020, Lechien, Chiesa-Estomba et al. 2020, Petrocelli, Cutrupi et al. 2021). Longitudinal studies are needed to further assess the relationship between OD and GD in COVID-19. Nevertheless, on average 4.8 months after infection and thus well after the acute phase, approximately 50% and 40% of patients reported persistent alterations in olfactory and gustatory function, respectively; these numbers are higher than what has been reported in some studies (Boscolo-Rizzo, Guida et al. 2021, Capelli and Gatti 2021, Lechien, Chiesa-Estomba et al. 2021) and lower than reported by others (Hopkins, Surda et al. 2021). The great variability in these results is due to very different study designs (self-report vs. psychophysical test; prospective vs cross-sectional) and studied populations (of different ethnicity and under different effects of selection bias), which either influence the measure of OD and GD in study populations or directly impact the baseline prevalence of OD and GD during COVID-19, offsetting all prevalence calculated at further points (Mazzatenta, Neri et al. 2020, von Bartheld, Hagen et al. 2020). For instance, in the study population included in this study, prevalence of OD decreases to 18.9% of participants when measured using the CPT at 4.8 (SD: 0.8) months after infection. The difference in these frequencies could be due to a higher sensitivity of the self-reported alterations compared to the CPT. Participants with milder forms of persistent hyposmia or with higher baseline olfactory sensitivity may have higher scores on the semi-objective CPT yet have not recovered entirely. We found a moderate-to-strong correlation between self-reported olfactory and gustatory changes, which were stronger than with self-reported trigeminal changes. This could be due to similar pathophysiological alterations in the olfactory and gustatory systems and their differences from that of the trigeminal system.

Knowing that the general population often mixes up retro-olfaction (perceiving odors from the substances in the mouth traveling posteriorly and rostrally to the olfactory epithelium) with taste, an alternative explanation would be a misunderstanding of this nuance by participants despite the fact that specific definitions for each modality were given (Landis, Frasnelli et al. 2005)(Malaty and Malaty 2013). The latter hypothesis is more probable since the correlation between gustatory self-report and CPT gustatory scores using strict gustatory stimuli (salt, sugar) is lower than the correlation between olfactory self-report and CPT olfactory scores. When tasting strictly gustatory stimuli in the CPT, participants reflect solely on their sense of taste, without the influence of retronasal sensations. These tests have the potential to be more accurate than simple subjective measures and could simplify large-scale psychophysical chemosensory testing. Others have reported the usefulness of similar self-administered chemosensory tests in the detection and follow-up of COVID-19-induced chemosensory dysfunctions *(Vaira, Salzano et al. 2020, Petrocelli, Cutrupi et al. 2021)*. Different theories have been proposed to explain the persistence of OD in certain individuals, ranging from olfactory epithelium dysfunction to central nervous system infection (Bilinska and Butowt 2020, Butowt and von Bartheld 2020, Solomon 2021). Since cells of the olfactory epithelium possess the ability to regenerate, the re-establishment of olfactory function is possible in the context of postinfectious OD (Cavazzana, Larsson et al. 2018), as well as in COVID-19-related OD, where 75-85% of the affected individuals recovered olfactory function within 60 days (Mullol, Alobid et al. 2020, Lechien, Chiesa-Estomba et al. 2021). The exact rate of olfactory recovery is still unknown, while post-COVID-19 OD prevalence ranging from 11%-60% at 6 months according to a recent study (Xydakis, Albers et al.). In addition to OD and GD, TD has also been reported in patients with COVID-19 (Cooper, Brann et al. 2020, Parma, Ohla et al. 2020).

Persistent chemosensory dysfunctions may be a sign of chronic central nervous system alterations (Gori, Leone et al. 2020, Wu, Xu et al. 2020), and there is now evidence that SARS-CoV-2 can infect olfactory sensory neurons in humans (Meinhardt, Radke et al. 2020, de Melo, Lazarini et al. 2021). Other viruses, such as the Japanese encephalitis virus, Varicella-Zoster virus, measles virus, human immunodeficiency virus and CoVs, were shown to invade the CNS (Koyuncu, Hogue et al. 2013). Febrile seizures, loss of consciousness, convulsions, ataxia, status epilepticus, encephalitis, myelitis, neuritis and extrapyramidal symptoms are among extra-pulmonary symptoms that have been described (Bohmwald, Gálvez et al. 2018). However, no evidence of intra-parenchymal replication has been found yet. Additional findings include the presence of local immune processes (Saussez, Sharma et al. 2021) and persistence of viral fragments in the olfactory epithelium (de Melo, Lazarini et al. 2021). Therefore, chronic post-COVID-19 inflammation in the olfactory pathway (epithelium, bulb) with or without direct infection is the most probable pathophysiological explanation of post-COVID-19 OD (Kirschenbaum, Imbach et al. 2020, Vaira, Hopkins et al. 2020, Xydakis, Albers et al.). The persistence of postinfectious neurological inflammation may contribute to the development or aggravation of chronic neurological diseases such as Parkinson, multiple sclerosis, or psychiatric outcomes (Morris 1985, Johnson-Lussenburg and Zheng 1987, Fazzini, Fleming et al. 1992, Murray, Brown et al. 1992, Stewart, Mounir et al. 1992, Cristallo, Gambaro et al. 1997, Arbour, Day et al. 2000, Koyuncu, Hogue et al. 2013, Cohen, Eichel et al. 2020, Taquet, Geddes et al. 2021). These patients should be followed up to document the development of neurological sequalae.

Moreover, approximately 10% reported parosmia and/or phantosmia following SARS-CoV-2 infection. These qualitative smell disorders usually involve unpleasant olfactory sensations (rotten eggs, sewage, smoke). While the exact patho-mechanism of parosmia and phantosmia are still to be elucidated, parosmia is probably linked to altered peripheral input/central processing of olfactory stimuli (Iannilli, Leopold et al. 2019). Importantly, patients with postviral OD and parosmia exhibit better recovery rates following olfactory training than those without parosmia (Liu, Sabha et al. 2021). Follow-ups will determine to what extent parosmia predicts a better outcome.

Women’s chemical senses were more affected than men during and after COVID-19 infection. Women typically have better scores in olfactory testing than men at baseline (Wang, Zhang et al. 2019). However, in line with our results, studies have revealed that women exhibit a higher prevalence and a longer persistence of postviral OD (Liu, Pinto et al. 2016, Sorokowski, Karwowski et al. 2019). Gender differences could be explained by a multitude of neuroendocrine, social, and cognitive factors (Sorokowski, Karwowski et al. 2019). We also found that older individuals have lower olfactory and gustatory sensitivities, especially during the acute phase of COVID-19.

Currently, there is no approved therapy specifically for COVID-19-induced OD, although experts agree that olfactory training could be prescribed for COVID-induced OD as it has a significant effect on olfactory function according to studies on other viral infections (Damm, Pikart et al. 2014, Sorokowska, Drechsler et al. 2017, Doty 2019, Huart, Philpott et al. 2021). Additionally, oral steroids, intranasal steroids and/or omega-3 supplements may be prescribed on an individual basis (Hopkins, Alanin et al. 2021). Most importantly, long-term follow-up of these patients will be necessary to assess other signs of neurological damage or spontaneous recovery, as recoveries can be possible after a year in other post-viral OD (Lee, Lee et al. 2014).

### Limitations

Given the cross-sectional design of the study, a recall bias is possible for all self-reported peri-SARS-CoV-2 infection values before or during the SARS-CoV-2 infection due to the 3-to-7-month gap. This study did not control for potential confounding factors like race and level of education. Finally, the CPT requires further validation for its gustatory and trigeminal components, and it relies on substances found in participants’ homes, which may lead to variation in test results due to the differences in the brand, quality, or expiration date of substances and consequently, their ability to trigger equal sensorineural responses.

## Conclusions

Nearly two thirds of SARS-CoV-2 infected patients had chemosensory impairments during their infection and despite improvements, impairments persist in half of them 3 to 7 months after COVID-19. Quantitative and qualitative olfactory dysfunction as well as persisting gustatory and trigeminal deficits were common in the cohort presented in this study. Given the frequency of these problems and the possible neurological underpinnings of these observations, it will be critical to understand the underlying mechanisms of these chemosensory dysfunctions, their evolution, and possible therapeutic options.

## Data Availability

Data can be made available upon request.

## Acknowledgments

We thank Josiane Rivard for preparing the online questionnaire, Cécilia Tremblay, Émilie Aubry-Lafontaine and Frédérique Roy-Côté for data collection and the validation of the Chemosensory Perception Test, and all study participants and frontline healthcare workers facing the COVID-19 pandemic. This work was supported by Fonds de recherche du Québec – Santé (chercheur boursier junior 2 #283144 to JF). NB and JF had full access to all the data in the study and take responsibility for the integrity of the data and the accuracy of the data analysis. FGL is the recipient of a tier-2 Canada research Chair. All authors declare no conflict of interest.

## Supplementary 1

### Validation of the Chemosensory Perception Test

This study was reviewed and approved by the research ethics board of the Université du Quebec à Trois-Rivières (CER-20-268-08-01.04). All participants provided a verbal or written informed consent prior to participation.

## Experiment 1

### Methods

Participants were recruited among previously tested groups. Olfactory testing was performed using the standardized Sniffin’ Sticks test at our laboratory from 2016 to 2019. Exclusion criteria was any perceived changes of their sense of smell since previous testing. Participants were distributed into 2 groups based on their Threshold-Discrimination-Identification (TDI) scores. The first group consists of participants with normal olfactory function (normosmia), defined as TDI scores above 30.5^61^. The second group had subjective olfactory dysfunction and equivalent TDI scores. Participants were administered CPT by means of a telephone interview.

### Results

TDI scores in the first group range from 32.5 to 41.5 (N= 19, 9 women and 10 men, age range [60-78]). TDI scores in the second group ranged from 8 to 30.25 (N= 17, 7 women and 10 men, age range [57-77]). CPT scores were positively correlated with the Sniffin’ Sticks (ρ=0.837, *P*<0.001). A cut-off score of 6 at the CPT had a sensitivity of 0.765 and specificity of 0.895.

## Experiment 2

### Methods

Participants were recruited among previously tested groups, participants from this cross-sectional study and in the public via social media. They were administered the University of Pennsylvania Smell Identification Test (UPSIT) (which was sent by mail) and the CPT under direct supervision through videoconferencing. Participants were distributed into 2 groups based on their UPSIT scores with a score equal or less than 33 in males and 34 in females defining hyposmia^62^.

### Results

UPSIT scores in the normosmic group ranged from 34 to 28 (n=29 (21 women), age range [22-73]). The hyposmic group had UPSIT scores ranging from 9 to 34 (n= 28 (21 women), age range: [22-72]). CPT scores were significantly correlated with the UPSIT score (ρ=0.377, *P*=0.004) in the whole group of participants. We found this correlation to be much stronger in the hyposmic group (ρ=0.702, *P*<0.001).

## Conclusions

The CPT allows for distinction between normosmia and olfactory dysfunction with high sensitivity and specificity. CPT scores are significantly correlated to UPSIT and Sniffin Sticks scores, especially in a group of individuals with olfactory dysfunction.

